# Development and Preclinical Evaluation of a CPAP-Compatible EtCO2 Sampler

**DOI:** 10.1101/2023.10.24.23297493

**Authors:** Sahil Sethi, Gene Hobbs, Devin Hubbard, Kenneth Donnelly, Joseph M. Grover, Imran Rizvi

**Affiliations:** Pritzker School of Medicine, University of Chicago, Chicago, IL 60637, USA; Joint Department of Biomedical Engineering, University of North Carolina at Chapel Hill, Chapel Hill, NC, North Carolina State University, Raleigh, NC 27599, USA; Department of Emergency Medicine, University of North Carolina, Chapel Hill, NC 27514, USA; Lineberger Comprehensive Cancer Center, School of Medicine, University of North Carolina at Chapel Hill, Chapel Hill, NC 27599, USA

**Author notes:** Corresponding authors (SS) (IR).

**Keywords:** End-tidal carbon dioxide, capnography, capnometry, oxygen therapy, noninvasive ventilation, continuous positive airway pressure, prehospital, emergency medical services

## Abstract

**BACKGROUND:** Capnography is a vital tool in EMS because changes to end-tidal CO_2_ (ETCO_2_) generally precede blood oxygen desaturation, and waveform morphology can be used to assess bronchial patency. Many of capnography’s indications overlap with those of continuous positive airway pressure (CPAP) therapy. However, there is currently no validated method to measure capnography during CPAP administration, and combining the two technologies in the prehospital setting lacks investigation in peer-reviewed literature.

**METHODS:** We developed a novel exhalatory CO_2_ sampler designed to seamlessly integrate with existing prehospital CPAP systems, manufactured via stereolithography. Using a Human Patient Simulator, we compared this novel sampler *in vitro* with a traditional nasal-oral cannula (NC), examining the effects of CPAP administration on ETCO_2_, capnogram shape, and respiration rate (RR).

**RESULTS:** The CCA maintained an air-tight seal during CPAP administration and showed no significant differences in ETCO_2_ values or RR compared to the NC. However, CPAP administration significantly reduced ETCO_2_ values and altered capnogram morphology for both samplers. These changes indicate that CPAP itself affects capnography readings, necessitating caution in interpreting these measurements.

**CONCLUSIONS:** The novel CCA is a viable tool for integrating capnography with CPAP in prehospital settings, but the effects of CPAP on capnography readings must be further studied. Future research should focus on clinical trials with human subjects to validate these findings and improve the interpretation of capnography data during CPAP therapy.

## Introduction

Respiratory issues are among the most common causes of 911 calls in the United States (1). Once on scene, EMS personnel interview the patient and use vital signs—such as heart rate, blood pressure, respiration rate (RR), pulse oximetry, and capnography—in addition to their physical exam findings to determine possible causes for their breathing difficulties. Capnography is particularly beneficial as it can typically detect changes to a patient’s breathing before pulse oximetry (2–4). For example, one study found that, for acute respiratory events, abnormal ETCO_2_ results were observed before SpO_2_ changes or observed hypoventilation in 70% of cases (2). Pulse oximetry is also known to overestimate oxygen saturation in patients with darker skin (5–7). Capnography’s physiological relevance stems from its correlation with PaCO_2_, which regulates the respiratory drive. Directly measuring PaCO_2_ requires an interventional test that is not practical in the prehospital setting, such as arterial blood gas analysis (8). There is some debate surrounding the correlation of ETCO_2_ to PaCO_2_, but several studies have found good correlation in healthy patients and patients with compromised pulmonary function who are spontaneously breathing (9–12). Capnography has been shown to be a valuable screening tool for mortality predictions in patients with sepsis (13,14). Also, in penetrating trauma patients, capnography measurements correlate with serum lactate levels (an indicator of tissue hypoxia and shock) and can predict the odds of the patient requiring operative intervention (15). See **Fig. 1B & 1D** for example capnograms.

**Figure 1.**
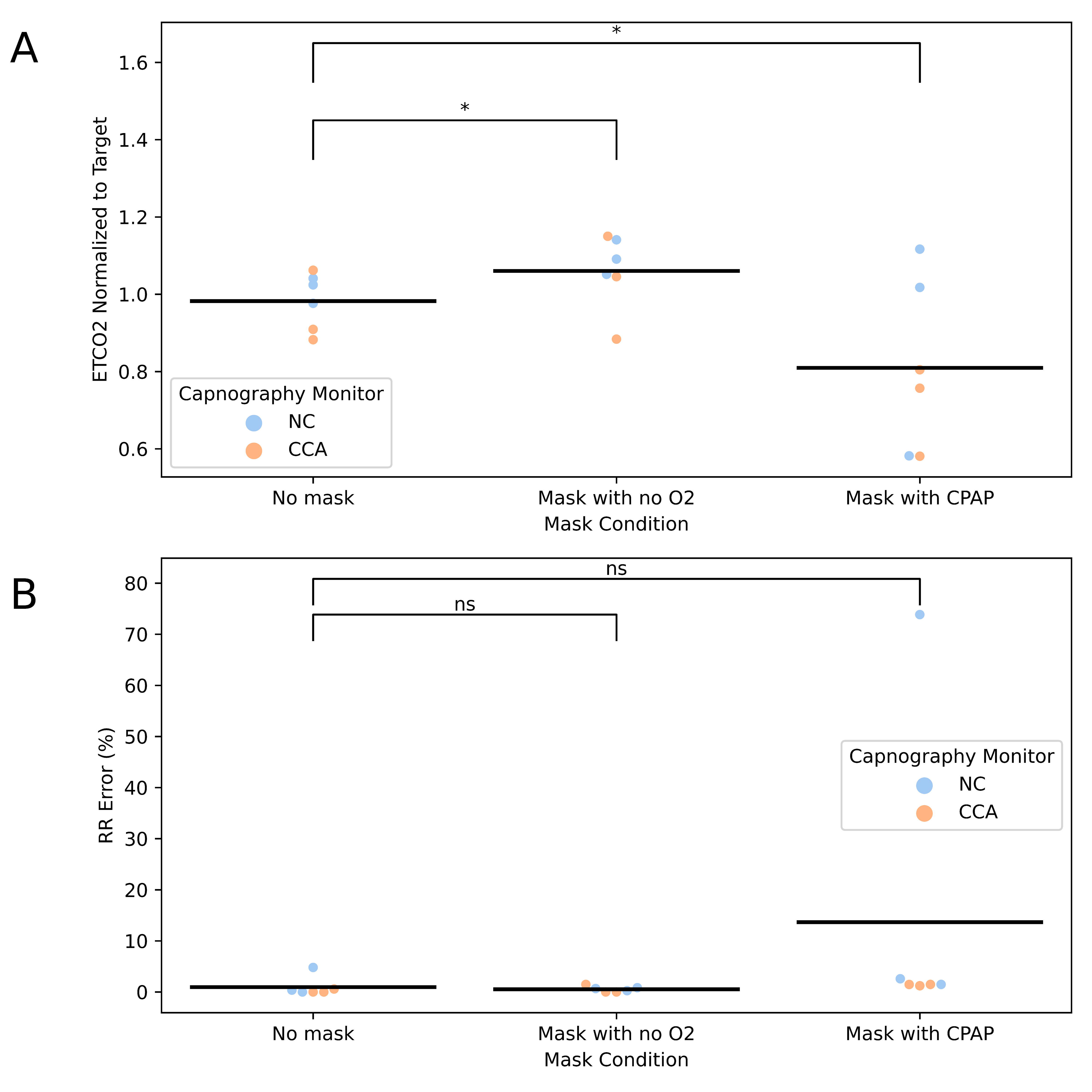
Capnography and Continuous Positive Airway Pressure (CPAP). (A) Nasal cannula (NC) commonly used for prehospital sidestream capnography sampling. (B) Example capnogram for a healthy individual (i) and an individual with bronchospasm (ii); peaks indicate exhalations while troughs indicate inhalations, and the end of the exhalatory plateau yields the PETCO_2_ measurement (labeled with a star). For the first waveform, phases I-IV are labeled, along with □- and □-angles. (C) Common prehospital CPAP setup with NC placed below the mask for capnography sampling; CO_2_ gas line is circled in purple. (D) Example EMS monitor depicting a capnography waveform; CO_2_ gas line insertion is circled in purple; standard orange capnography port (not shown) is used for insertion.

Prehospital capnography is most commonly used for validating endotracheal tube placement (16), as well as helping identify asthma, chronic obstructive pulmonary disease (COPD), pulmonary embolism (PE), and congestive heart failure (CHF) (17–19). The morphology of the capnogram is crucial for assessing bronchial patency as bronchospasm tends to yield a “shark fin” waveform (prolonged phase II with increase or loss of L-angle) (20,21), which is depicted in **Fig 1B.ii**. Monitoring of the waveform is currently used to assess response to bronchodilators (20,22). Prehospital indications for CPAP strongly align with those of capnography and include CHF, COPD, asthma, acute bronchitis, pneumonia, and near-drownings with signs of pulmonary edema (23,24). Prehospital CPAP has been shown to improve patient outcomes by reducing the need for intubation (and thus decreasing the likelihood of developing ventilator-associated pneumonia) (25), improving vital signs, and reducing myocardial damage—its use is also associated with reduced mortality (24,26). Using capnography to monitor the effectiveness of CPAP would allow for the rapid determination of ineffective therapy and a quick decision to be made regarding switching to an invasive option (27,28). This would also align with the 2011 AARC Clinical Practice Guideline on capnography, which recommends its use for monitoring response to therapies that improve the ventilation-perfusion (V/Q) ratio (23).

Despite these points, the two technologies are not currently easily compatible. Common EMS cardiac monitors with built-in sidestream capnography sensors, such as the Physio-Control LIFEPAK-15, are used with a nasal-oral cannula (NC) sampler for spontaneously breathing patients, and an in-line sampler for intubated patients (27,29). Non-invasive ventilation (NIV) dilutes exhalations, so sampling as close to the nares/mouth as possible is best (27). Thus, using an NC sampler is generally preferred during NIV (30). However, placing a cannula beneath a CPAP mask is not ideal as it allows gas to escape at the location where the sampling line exits the mask (27); significant leaks are problematic as they decrease CO_2_ elimination and ETCO_2_ values (30). Despite this, due to anecdotal evidence, some EMS education sources recommend using the NC sampler for capnography during CPAP (31–33), and EMS systems have already added this recommendation to their protocols (34).

In this study, we aim to design and manufacture a novel exhalatory CO_2_ sampler, referred to as the CPAP-Capnography Adapter (CCA), that will be compatible with existing prehospital CPAP devices. The CCA will be developed to maintain the system’s airflow while ensuring compatibility with existing EMS cardiac monitors that feature capnography monitoring capabilities. To assess the performance of this novel device, we will conduct preliminary *in vitro* testing using a Human Patient Simulator (HPS). This testing will compare the CCA to the traditional nasal-oral cannula (NC) to evaluate the impact of CPAP administration on ETCO2 measurements, capnogram morphology, and respiratory rate (RR).

## Materials and Methods

### CPAP-Capnography Adapter (CCA) Design Process & Manufacturing

A novel method for measuring capnography during CPAP was invented and tested alongside the nasal-oral cannula method. Engineering sketches are provided in **Fig. 2**. The new method involved attaching an elbow-shaped adapter to a prehospital CPAP setup between the mask and flow regulator. To ensure an air-tight seal, flat ring seals were created out of duct tape and applied to each end of the elbow (functioning similarly to square profile O-rings). A nasal sampling element was fed through this adapter and into the mask, allowing for sidestream capnography sampling. To prevent the nasal sampler’s tubing from getting crimped by the elbow as it passed into it, a short internal guide tube was added to provide greater support. The nasal sampler base slid onto existing CapnoLine® tubing containing a standard orange capnography plug; the sampler fit snugly via friction but was further secured via duct tape. Combined, this setup is referred to as the CPAP-Capnography Adapter (CCA).

**Figure 2.**
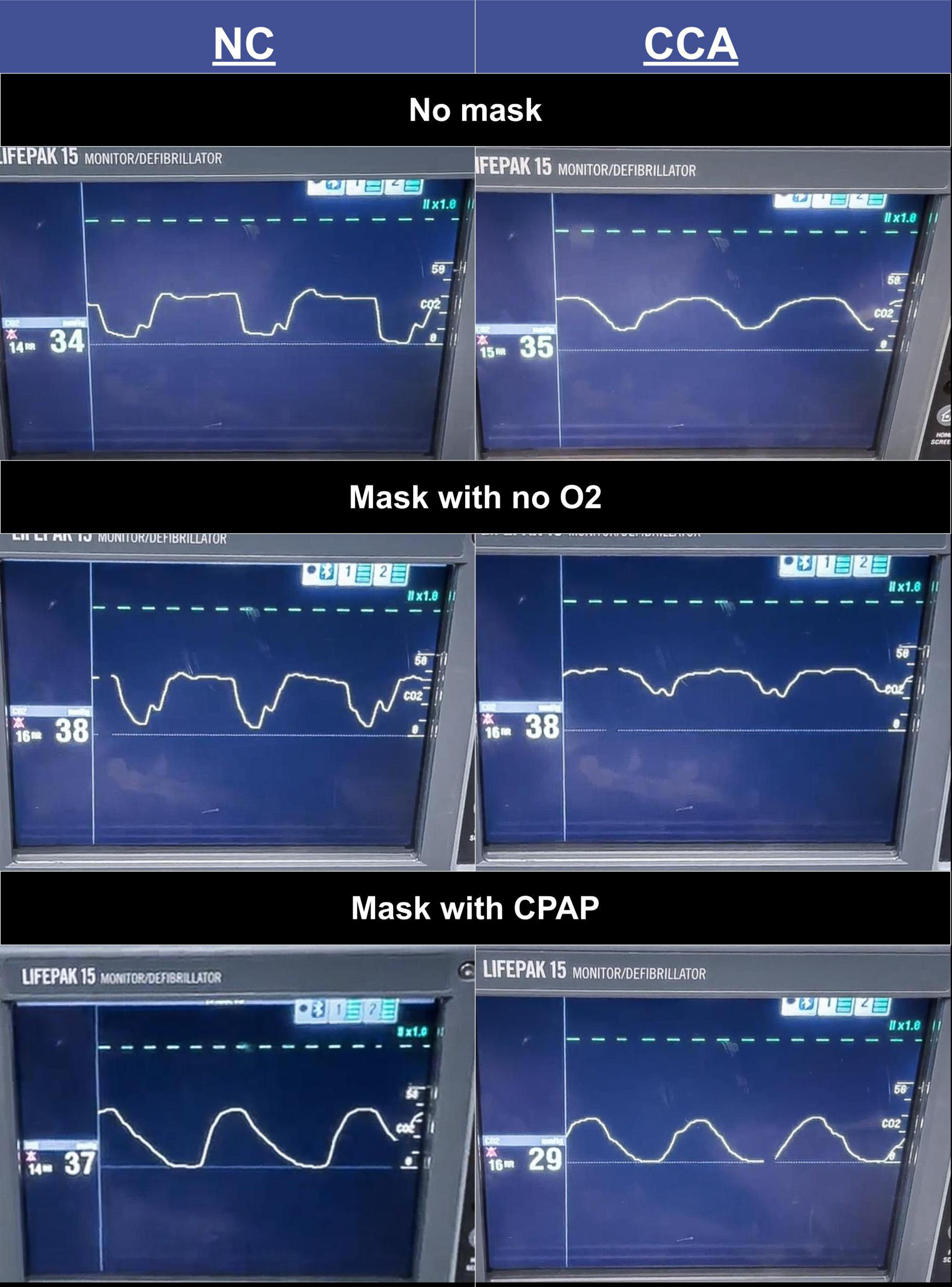
Design sketches for novel CPAP-capnography adapter (CCA) system. Orthographic and isometric views are depicted for both the elbow adapter (top) and nasal sampler (bottom). All dimensions are in millimeters, and angles are in degrees. These sketches represent the conceptual design of the CCA.

The CCA elbow and nasal sampler were designed using OnShape Computer-Aided Design (CAD) software and printed with a Formlabs Form 2 stereolithography (SLA) 3D printer (see **Fig. 3A**). Formlabs Clear V4 resin was selected as the elbow material due to its translucency and rigidity (35). Formlabs Elastic 50A resin was selected as the nasal sampler material due to its translucency and flexibility (36), the latter property allowing for the accommodation of various nose sizes. After printing, an isopropyl alcohol (IPA) bath was used to vigorously wash parts for 15 minutes. Then, an IPA-filled syringe was used to clear excess resin from small channels, and the parts were soaked in a clean IPA bath for an additional 15 minutes. Next, an ultraviolet (UV) light & heat cure was conducted according to the manufacturer’s guidelines (power density: 1.25 mW/cm^2^; LED centered at 405 nm; 60 °C for 20 minutes for Elastic 50A and 60 minutes for Clear V4) (35–37).

**Figure 3.**
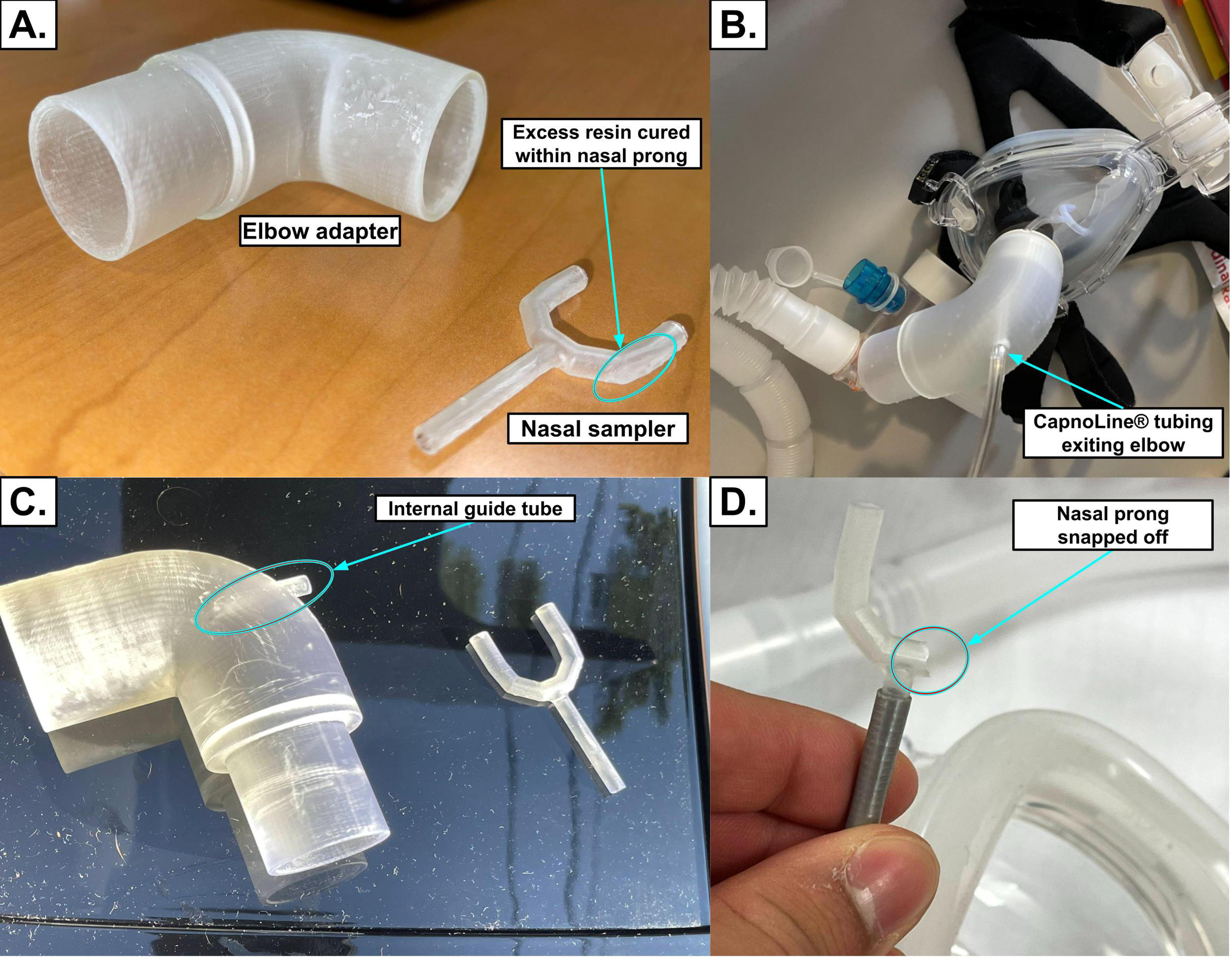
CPAP-Capnography Adapter (CCA) manufacturing & experimental setup. (A) The CCA being manufactured using the stereolithography 3D printing process. (B) A labeled diagram of the final CCA design. (C) Integration of the CCA into a prehospital CPAP setup. (D) A display from the Human Patient Simulator (HPS) showing modeled ETCO_2_ & respiration rate. (E) CCA applied to the HPS without a CPAP mask. (F) CCA integrated into a prehospital CPAP setup and applied to the HPS.

### Materials

A stock CPAP kit containing a Pulmodyne O_2_-MAX™ and Curaplex O_2_ Max Trio BiTrac ED Mask was used. For all experimental groups, CPAP tubing was fully contracted to standardize dead space, and the PEEP valve was set to 10 cm H_2_O. The following nasal-oral cannula was used: Covidien Smart CapnoLine® Plus O_2_, Adult/Intermediate CO_2_ Oral/Nasal Sampling Set with O_2_ Tubing. For their respective trials, the CCA and NC were plugged into the capnography port on a Physio-Control LIFEPAK-15 monitor to obtain capnograms and ETCO_2_ values. As the HPS is meant to accurately model a variety of respiratory parameters, ETCO_2_ and respiration rate could not be directly manipulated to test the full range of expected values for each because this could result in a biologically inaccurate combination of parameters. Therefore, pre-programmed patient respiratory profiles were used for testing. For each experimental group, the HPS was kept in the supine position, and CCA and NC positioning were kept constant.

### Human Patient Simulator

The CAE Human Patient Simulator (HPS) is a high-fidelity medical training tool designed to replicate human physiological responses, with a particular emphasis on modeling respiratory functions. It features realistic chest rise and fall, variable lung compliance and resistance, and the ability to simulate spontaneous breathing, apnea, and mechanical ventilation scenarios. Vital signs can be measured on it with traditional tools, as it exhales oxygen and inhales carbon dioxide; blood pressure can also be manually auscultated (38). These capabilities make it an incredibly useful tool for preclinical testing of novel medical devices, such as in this study. Even for low-risk devices such as the CCA, such preclinical testing is essential to determine if the new device warrants the time and cost of conducting human trials.

### Experimental Design

The following six configurations were tested: (1) NC with no CPAP mask nor O_2_ flowing, (2) CCA with no CPAP mask nor O_2_ flowing, (3) NC with CPAP mask but no O_2_ flowing, (4) CCA with CPAP mask but no O_2_ flowing, (5) NC with CPAP mask and O_2_ flowing, and (6) CCA with CPAP mask and O_2_ flowing. Groups 1 and 2 are controls for evaluating the effects of CPAP mask dead space (Groups 3 & 4) and CPAP therapy (Groups 5 & 6) on capnography sampled via NC and CCA, respectively. In groups 3 and 4, the CPAP mask was applied, but the tubing was not connected to an oxygen source and remained open to ambient air, allowing the mannequin to breathe room air. This setup was included to assess the effect of added anatomic/mechanical dead space from the CPAP mask and tubing on capnography measurements, as previous studies have shown that simply applying a mask can increase ETCO_2_ (39). These groups served as positive controls, validating that our setup can detect the changes in ETCO_2_ that have been observed in other studies, thereby allowing us to better contextualize the results from the other experimental groups where CPAP was actively delivering oxygen. All experimental groups are summarized in **Table 1**.

**Table 1.**
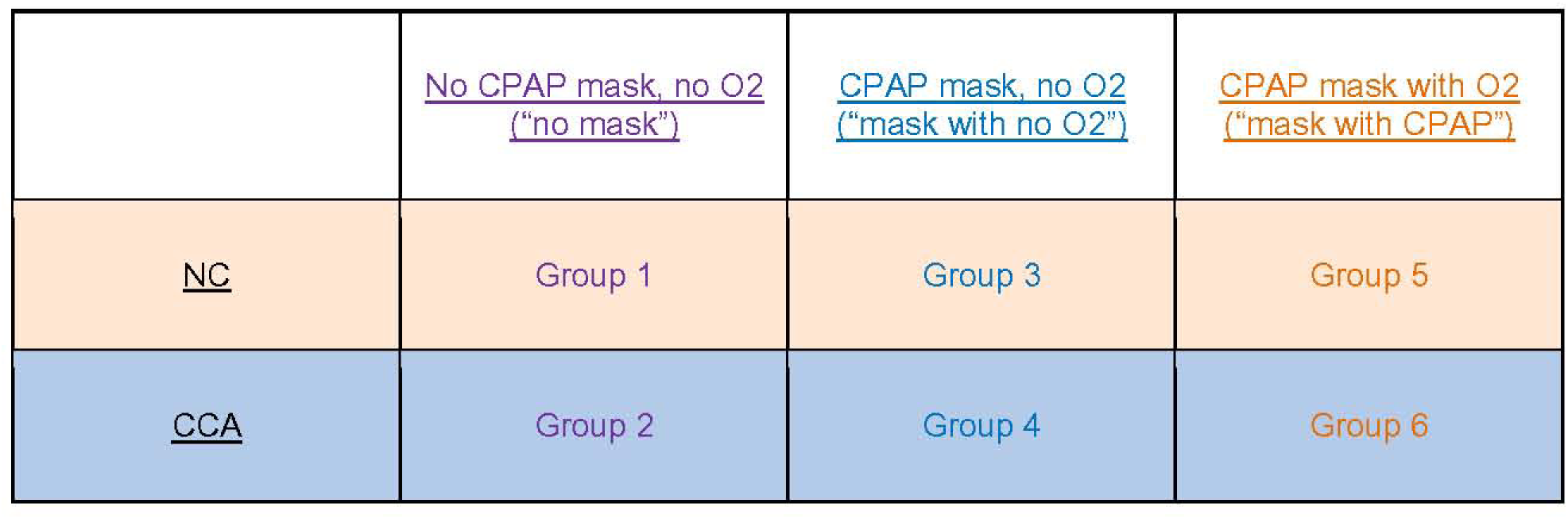
Experimental groups used for testing on Human Patient Simulator (HPS). Groups 1-6 were each tested on three HPS standardized simulated patients: Stan D. Ardman II (Healthy Adult Male), Grandma Smokey (COPD Patient), & Kenton Parkers (CHF & COPD Patient); these profiles have set-point ETCO_2_ values of 33 mmHg, 22 mmHg, and 34 mmHg, respectively.

For groups 1-6, data were obtained from three standardized simulated patients from the HPS system: Stan D. Ardman II (Healthy Adult Male), Grandma Smokey (COPD Patient), & Kenton Parkers (CHF & COPD Patient). From their pre-programmed respiratory profiles, their ETCO_2_ targets were 33 mmHg, 22 mmHg, and 34 mmHg, respectively. The HPS continuously adjusts modeled vital signs to remain consistent with this ETCO_2_, as well as the rest of its pre-programmed patient profile. As groups 1-6 were tested for all three patients, 18 total trials were conducted. For each trial, the ETCO_2_ value and respiration rate were recorded from the LIFEPAK-15 monitor’s screen for 20 consecutive breaths. Software limitations of the LIFEPAK- 15 and HPS prevented the direct export of the data to a spreadsheet. Consequently, videos were taken of both displays, timestamps were lined up, and values were entered into a spreadsheet to facilitate data collection. Trials were spaced one minute apart to allow for the modeled vital signs from the HPS to stabilize in response to changes. Only 20 breaths per group were measured because the HPS is designed to maintain constant respiratory profiles, resulting in minimal variation once values have stabilized. Measuring more than 20 breaths within a single trial would not provide additional variability but could lead to oversampling, artificially inflating the precision of the data. Additionally, this method aligns with a similar study that used an ASL-5000 lung simulator to compare various methods of interfacing capnography with hospital oxygen masks (39). By averaging data across 20 breaths, we reduce the risk of overfitting while capturing representative data from each trial.

### Statistical Analysis

For all tests, alpha was chosen to be 0.05. As each simulated patient had a different set-point ETCO_2_, measured ETCO_2_ values at each breath were normalized by dividing by their respective target ETCO_2_. Unlike ETCO_2_, the HPS displayed the true RR value for each breath, so the true and measured values were used to calculate RR percent error. Values were averaged across the 20 breaths in each trial to avoid overfitting during analysis.

Normalized ETCO_2_ and RR percent error were analyzed for any effects caused by the following nominal independent variables: capnography sampler (NC or CCA), mask condition (no mask, mask with no O_2_, and mask with CPAP), and patient (Stan D. Ardman II, Grandma Smokey, & Kenton Parkers). In accordance with the current literature (40–42), a mixed effects model was constructed with “patient” as a random effect. Using the mixed model (multilevel linear model) feature within JMP Pro 17, the full factorial of capnography sampler, mask condition, and patient were selected as model effects; all terms containing “patient” were marked as random effects. The t-statistics were then used to obtain specific p-values. All statistics were also validated using Python’s statsmodels package.

## Results

### Development of CPAP-Capnography Adapter

The first prototype of the CPAP-Capnography Adapter (CCA) is depicted in **Fig. S1A**. An elbow-shaped design was chosen to allow the CapnoLine® tubing to pass through a hole in the wall of the adapter without needing to bend—which could cut off gas flow (see **Fig. S1B**). To ensure compatibility with existing EMS cardiac monitors and ensure the final CCA is simple and cheap to manufacture, existing CapnoLine® tubing with a standard orange capnography port plug was used. For this prototype, both components were printed with Clear V4 Formlabs resin, allowing them to be manufactured together in a single print.

Despite precisely measuring the dimensions of the existing CPAP setup, the first prototype CCA did not have an air-tight fit with the mask; air could be both felt and heard escaping. Further, the existing CapnoLine® tubing could get easily crimped if the mask was moved around and the tubing not kept perfectly straight. Additionally, despite soaking and vigorously washing the elbow adapter and nasal sampler in isopropyl alcohol (IPA) per Formlabs’ guidelines prior to the UV (ultraviolet light) bath, excess resin was found within one of the prongs of the nasal sampler, narrowing its internal diameter on that side. As a result, several changes were implemented for the second prototype.

The second prototype added an internal guide tube to better distribute shear forces on the CapnoLine® tubing, reducing the likelihood of crimping (see **Fig. S1C**). Next, a syringe was filled with IPA and used to flush out the channels within the nasal sampler of any excess resin. Last, flat ring seals were created out of duct tape and applied to each end of the elbow (functioning similarly to square profile O-rings) (see **Fig. 3B-C**).

The adjustments to the second prototype were all successful—the internal guide tube prevented crimping upon movement of the CPAP mask, the nasal sampler was free of excess resin, and no leaks could be heard or felt upon connecting the CCA to a CPAP mask and turning on the oxygen supply. However, the rigidity of the nasal sampler caused it to snap on several occasions when removing the 3D printing supports and sanding it down. The rigid material might also be uncomfortable to a human patient, so the decision was made to switch to Elastic 50A resin for the nasal sampler only. While this required an additional print, the small size of the nasal sampler ensured that this did not add to the complexity of the manufacturing process.

Three 3D prints of the final design were created to ensure reproducibility (though only one was tested on the HPS to ensure standardization). Supports were auto-generated for each print, causing a slightly different total resin amount from print to print. However, the elbow adapter remained below 40 mL resin per print, and the nasal sampler remained below 5 mL resin per print. Thus, the total cost of the CCA resin remained below $7 (based on current resin prices of $149/L for Clear V4, and $199/L for Elastic 50A) (43). The remaining parts of the setup, such as the CapnoLine® tubing/plug and tape, are commonly found in ambulances that carry monitors with capnography monitoring capabilities.

### Effects on Capnography Waveform

For all three patients during the experiment, it was noted that a leak was audible and could be felt by the experimenter’s hands around the sides of the CPAP mask for the NC groups when oxygen was administered (Group 5); no such leaks were noticed for the corresponding CCA group (Group 6).

Representative capnography waveforms are depicted in **Fig. S2**. Within each trial, waveform shape remained consistent across the 20 recorded breaths. It was noted that the waveforms obtained via the CCA appeared to consistently show more dead space compared to the NC capnograms, indicated by the waveform not fully returning to 0 mmHg during each inhalation. This was most noticeable in the “no mask” and “mask with no O_2_” conditions. These same capnograms appeared to have increased □- and □-angles compared to those from the NC, with bilateral blunting of the waveforms.

Across both the NC and CCA trials, waveform shape remained similar when transitioning from the “no mask” to the “mask with no O_2_” condition, but a small increase in dead space was noted. Note that the CPAP setup allowed for exhaled gas to escape through a valve near the PEEP regulator when oxygen was not flowing, reducing CO_2_ rebreathing; the high flow rate during CPAP forces this valve closed to prevent leaks while administering the therapy. The “mask with CPAP” waveform consistently had a lower magnitude and sharper peak compared to the “no mask” and “mask with no O_2_” conditions. Specifically, the expiratory upstroke (phase II) appeared relatively unaffected, but the alveolar plateau (phase III) and inspiratory downstroke (phase IV/0) were severely blunted, with L-angles appearing to be around 180 degrees. These morphological changes were present when administering CPAP in all three patients. There were no noticeable consistent differences between NC and CCA waveforms when CPAP was being administered.

### Effects on End-Tidal CO2

Results for “ETCO_2_ normalized to patient” compared against “capnography sampler” and “mask condition” are depicted in **Table 2** and **Fig. 4A**. Capnography sampler was not found to significantly affect normalized ETCO_2_ (NC: 1.005, CCA: 0.897; p = 0.25). The interaction between mask condition and capnography sampler was also found to have no significant effect on normalized ETCO_2_ (p = 0.69), but mask condition alone did have a significant effect (p = 0.031). Specific model parameter t-statistics revealed that the “mask with CPAP” normalized ETCO_2_ was significantly less than that of the “no mask” condition (0.810 vs. 0.983, respectively; p = 0.015); moreover, the “mask with no O_2_” normalized ETCO_2_ was significantly greater than that of the “no mask” condition (1.060 vs. 0.983, respectively; p = 0.033).

**Table 2.**
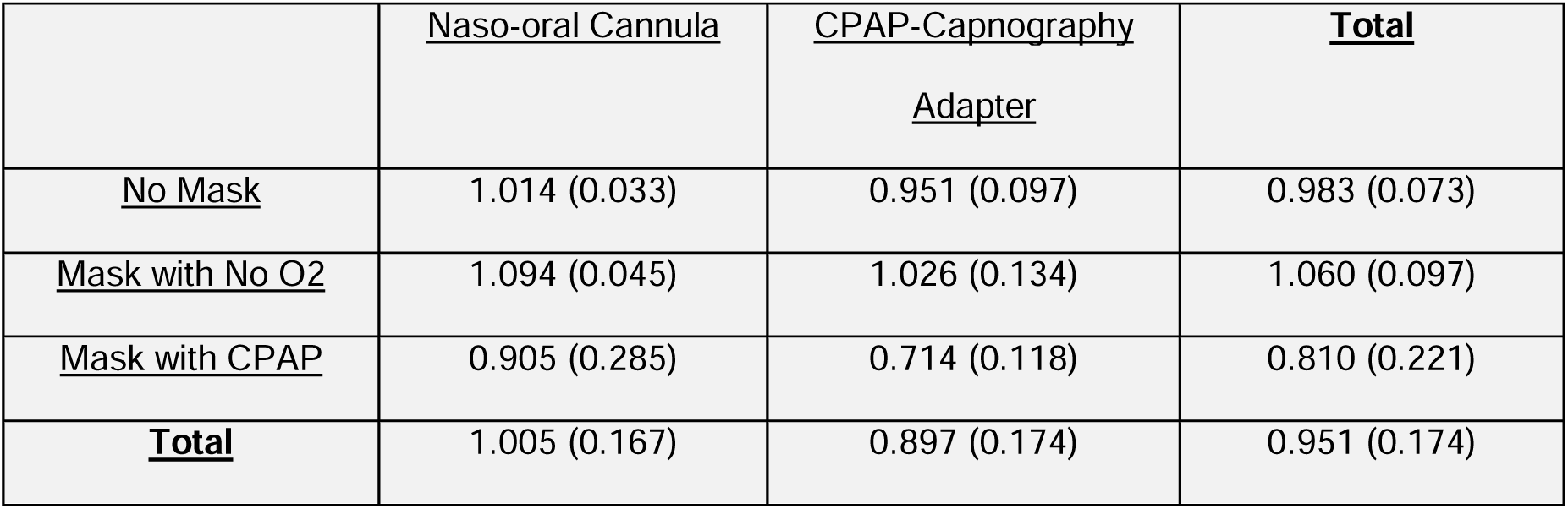
End-tidal CO_2_ (ETCO_2_) normalized to target for all experimental groups. Standard deviation is provided for each respective value in parenthesis immediately after it.

**Figure 4.**
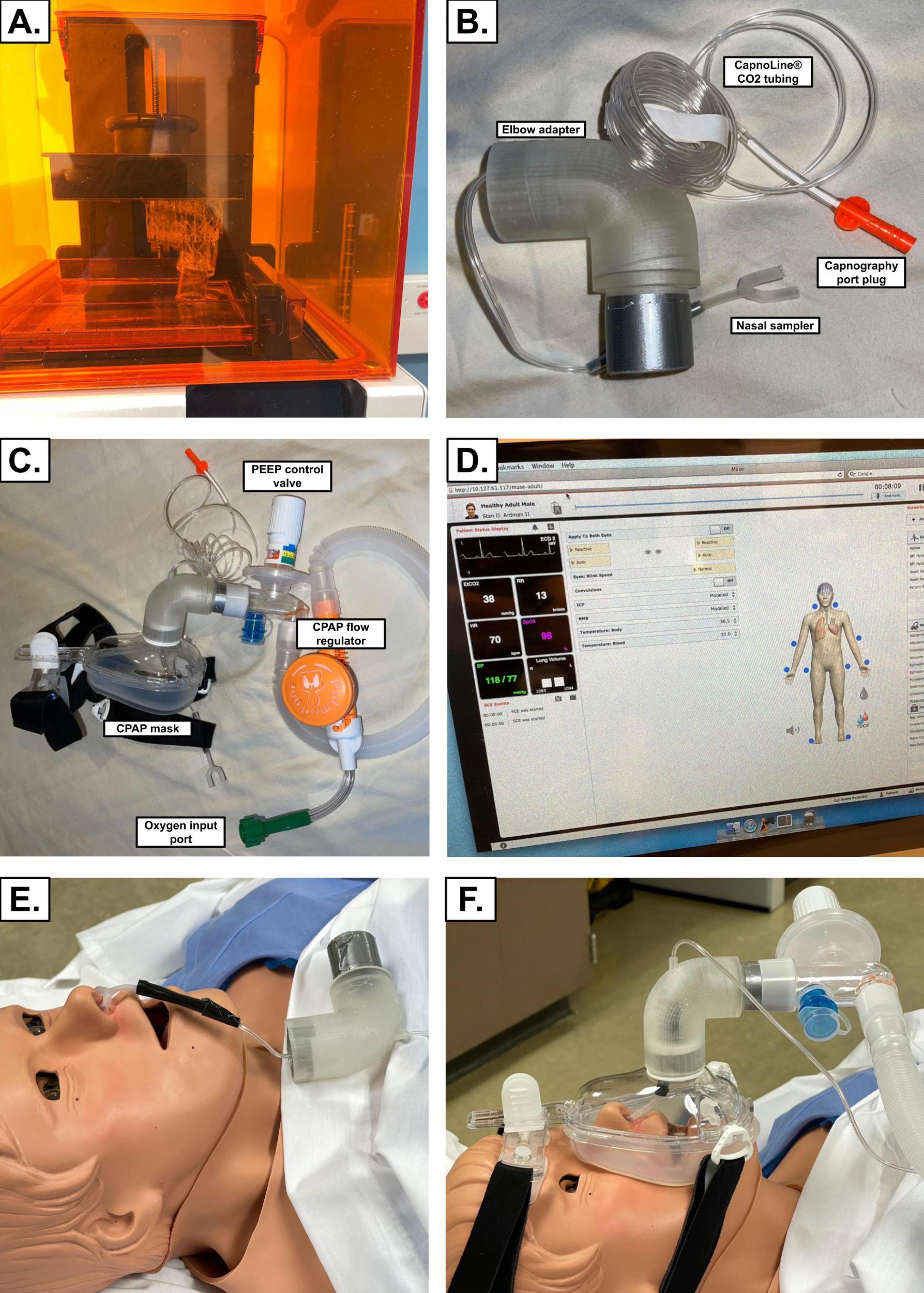
Normalized ETCO_2_ and RR percent error plotted by mask condition. Horizontal black lines indicate average values for each mask condition. No significant differences were found between NC (naso-oral cannula) and CCA (CPAP-capnography adapter) groups, so both are shown together. * indicates p < 0.05 from the mixed-effects model. Data points reflect the 20-breath average per trial. CPAP = continuous positive airway pressure, O_2_ = oxygen gas.

### Effects on Respiration Rate

Respiration rate percent error data based on mask condition and capnography sampler are depicted in **Table 3** and **Fig. 4B**. Capnography sampler did not significantly affect RR percent error (NC: 9.423%, CCA: 0.690%; p = 0.40). Mask condition and the interaction between mask condition and capnography sampler were both found to have no significant effect on RR percent error (p = 0.37 & 0.44, respectively). One of the 20-breath trials produced a high average RR percent error for the ‘mask with CPAP’ and ‘NC’ conditions (13.677% & 9.423%, respectively). In the Group 5 (NC; mask with CPAP) condition for the patient ‘Kenton Parkers’ (CHF & COPD Patient), an artifact consistently caused the LIFEPAK-15 monitor to double the measured respiration rate for all 20 breaths. This artifact reflects a potential limitation of the NC when used with high-flow CPAP. While this artifact increased the mean and variance in the Group 5 data, removing this trial did not alter the overall results. The statistical analysis showed no significant differences in RR percent error between mask conditions or capnography samplers whether the artifact was included or not. Therefore, we retained this trial in the analysis to highlight a potential limitation of the NC during high-flow CPAP.

**Table 3.**
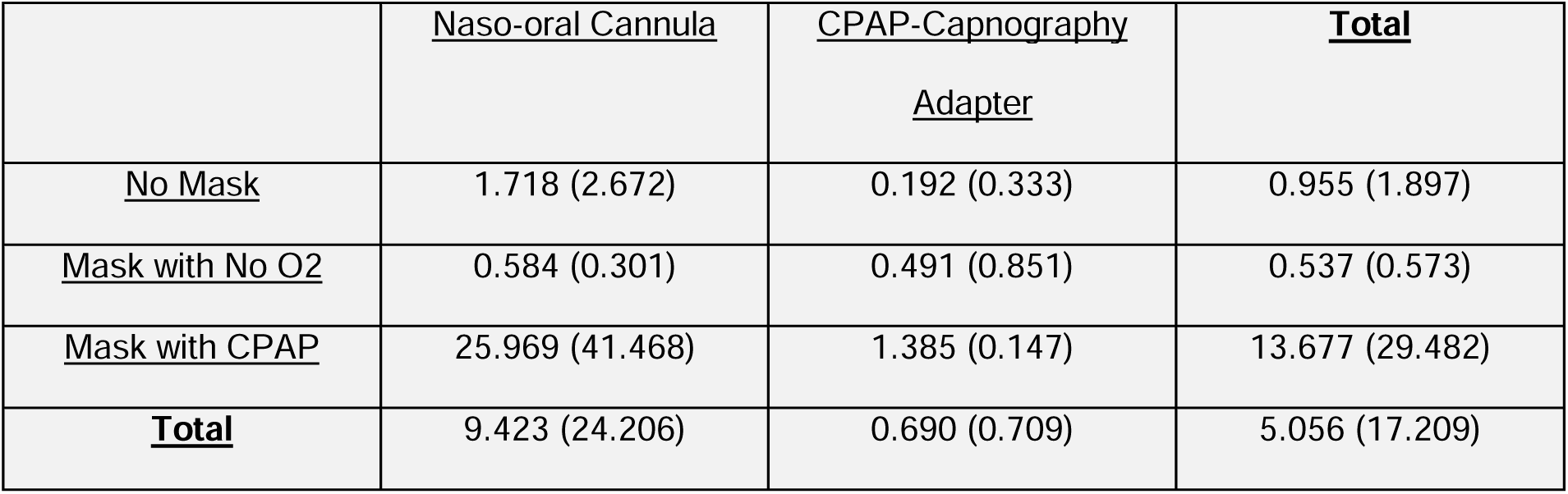
Respiration rate (RR) percent error for all experimental groups. Standard deviation is provided for each respective value in parenthesis immediately after it.

## Discussion

### Development of CPAP-Capnography Adapter

The final design of the CCA met its design objectives: it is compatible with existing EMS cardiac monitors (tested via the LIFEPAK-15) and does not cause any identifiable leaks during CPAP administration. Further, the low cost of manufacturing ensures that expense would not be a significant barrier to the adoption of the device in the clinical setting. However, data from human subjects would be necessary prior to such adoption, which is outside the scope of this study. Initial preclinical evaluation, as conducted here, is essential to determine if the device warrants the time, resources, and ethical considerations of conducting human studies. High-fidelity models such as the HPS used here are useful tools for such preclinical evaluation.

### Effects on Capnography Waveform

The main difference between the NC and CCA waveforms was the observed increase in dead space—indicated by the capnogram not returning to baseline during inhalation—and altered morphology when using the CCA. This likely resulted from CO_2_ accumulating in the sampling line, though it did not trend upward, indicating that exhalations were able to escape to some extent. The increased CO_2_ within the mask observed here is consistent with increased mechanical/anatomic dead space—a known factor when using any breathing apparatus that introduces additional space between the patient’s airway and the point of exhalation (44). While the increase in dead space is consistent with previous studies, such as Phillips et al. (2017) (39), further testing is needed to confirm which component of the CCA’s design caused this artifact. Specifically, a Computational Fluid Dynamics (CFD) model could be used to analyze differences in gas flow between the NC and CCA during CPAP administration. Importantly, the CCA is intended for use specifically during CPAP, and it produced nearly identical waveforms to the NC in the CPAP trials. Therefore, the altered morphology of the CCA waveforms in the “no mask” and “mask with no O_2_” conditions has limited clinical impact, as these conditions are not representative of typical CPAP usage.

However, the fact that CPAP consistently blunted the capnograms from both the NC and CCA is more concerning. The ETCO_2_ value and capnogram morphology are critical in identifying causes of breathing difficulties and monitoring responses to therapy. For example, the phase III alveolar plateau slope is an indirect marker of ventilation-perfusion (V/Q) mismatch, and changes in the α- and β-angles of the capnogram can suggest bronchospasm, airway obstruction, or rebreathing (21). During CPAP, these waveform features are altered or diminished, limiting the utility of capnography in identifying these respiratory conditions. Therefore, prehospital care providers must exercise caution when interpreting waveform morphology during CPAP, as the typical diagnostic features may not be reliably visible. Future research should investigate how waveform morphology changes under CPAP in both healthy individuals and patients with respiratory conditions to better understand how capnography can be used in this setting.

### Capnography Sampler Effect on ETCO_2_ and Respiration Rate

Overall, no differences were found between ETCO_2_ and RR percent error when measuring capnography via a traditional nasal-oral cannula (NC) or the novel CPAP- Capnography Adapter (CCA). The observation that a leak between the face and mask was present during CPAP in all NC trials and absent in all CCA trials indicates that the CCA met its design objectives. While the NC did have a higher RR error than the CCA (9.423% vs. 0.690%), this can be attributed to a single trial (Group 5 for the patient “Kenton Parkers”) where an artifact caused the LIFEPAK-15 to record a doubled respiration rate. This artifact was not noted in any other trials, and its effect was not statistically significant. However, this error was present for all 20 breaths of the trial in question, so it should not be ignored; it is possible that something about the NC or particular respiratory profile used for the trial predisposed the monitor to make the error. Additional experiments would be needed to determine if the artifact is truly associated with the use of the NC or if it originates from another source.

### Mask Condition Effect on ETCO_2_ and Respiration Rate

Administering CPAP significantly reduced the measured ETCO_2_ (0.810) when compared to “no mask” (0.983), which is consistent with the waveform observations. While a reduction in ETCO_2_ could indicate reduced PaCO_2_, the fact that the HPS did not model ETCO_2_ and instead had a set target value indicates that the measured ETCO_2_ did not accurately reflect the mannequin’s breathing profile during CPAP. Both capnography samplers recorded ETCO_2_ values within five percent of the HPS’s target ETCO_2_ in the control “no mask” condition (NC: 1.4% higher; CCA: 4.89% lower), so their measurement of the partial pressure of CO_2_ at the HPS’ nares was relatively accurate. Consequently, the notable decrease in ETCO_2_ during CPAP indicates that the CO_2_ at the nares was diluted by the high oxygen flow rate (which can exceed 100 L/min) (45). Thus, the reduction in measured ETCO_2_ is not an artifact of the capnography samplers themselves. As a result, this ETCO_2_ likely would no longer correlate with PaCO_2_ in a human subject. Further evidence that the functionality of the capnography samplers is not compromised by CPAP is the fact that no differences in RR percent error were found between the “no mask”, “mask with no O_2_”, and “mask with CPAP” conditions—indicating that the capnography samplers obtain sufficient gas samples to record measurements during all mask conditions. Therefore, healthcare professionals should use ETCO_2_ values obtained during CPAP with caution, as a decreased ETCO_2_ value might not indicate a true decrease in a patient’s PaCO_2_.

### Relationship with Previous Studies

In 2012, Razi et al. showed good correlation between PaCO_2_ and ETCO_2_ obtained during mechanical ventilation (including CPAP) in a hospital setting. However, these patients were intubated and monitored in a mainstream manner (46). In 2015, Piquilloud et al. used an NC sampler to monitor capnography in ICU patients receiving non-invasive ventilation (NIV), and found that ETCO_2_ was inadequate to predict both PaCO_2_ value and variations over time. However, they only included 11 patients in their study, and used bi-level positive airway pressure (BiPAP) instead of CPAP (47). A much larger study of 154 patients was published in 2021 by Uzunay et al. that showed a significant difference between ETCO_2_ and PaCO_2_ during both non-invasive BiPAP and CPAP in the emergency department. However, a positive moderate correlation was present, so the authors stated that capnography monitoring during NIV may still be beneficial for predicting PaCO_2_ changes in response to therapy—allowing for a quicker decision to be made regarding switching to intubation (28). The mainstream capnography monitoring method they used is not compatible with prehospital CPAP nor existing EMS cardiac monitors, though. Most recently, Sakuraya et al. showed in 2022 that PaCO_2_ and ETCO_2_ correlate well during CPAP when measured via an NC in a sidestream manner, and correlate even better when measured in a mainstream manner. Their study was conducted on 60 recently extubated ICU patients. The authors concluded that the results of previous studies monitoring ETCO_2_ during NIV were mixed likely due to physiological issues and shortcomings of exhalation sample collection (12).

None of the existing studies described changes to waveform morphology nor used a prehospital CPAP system. Further, Sakuraya et al. is the only one that used a capnography sampler compatible with common EMS cardiac monitors during non-invasive CPAP. All the studies presented here assessed the correlation of PaCO_2_ to ETCO_2_ in human subjects, and they achieved mixed results likely due to physiological factors and variations in methodology. Our study complements this work by showing that—with physiological factors and variation removed through the use of an HPS—ETCO_2_ value and capnogram morphology are directly compromised by CPAP itself. We could not assess PaCO_2_ due to the limitations of the mannequin used here; however, we show that the ETCO_2_ value and capnogram shape should not be interpreted with current protocols. Additional research into interpreting the value and waveform when taken during CPAP is needed.

### Limitations

The primary limitation of this study is that it was conducted using simulated patient profiles on a Human Patient Simulator (HPS). The CAE Healthcare HPS is extremely accurate at mixing respiratory gasses and mimicking human breathing, so it is commonly used in anesthesia training (38,48–50). Using an HPS rather than human subjects allows for greater control of confounding variables, facilitates rapid data collection, and relieves the ethical concerns (and IRB approval) of testing a novel capnography sampler on patients in respiratory distress who are receiving CPAP. However, it is not a perfect substitute for testing on human subjects. For example, human patients exhibit significant differences in lung compliance, airway dynamics, and facial anatomy, all of which can affect the seal of the CPAP mask and, consequently, the measurement of CO_2_. Mask leakage, in particular, may behave differently in human patients than in the HPS, which could influence ETCO_2_ measurements and waveform morphology.

The HPS used in this study also has fixed physiological parameters, which limits its ability to simulate real-time variability seen in human patients. In clinical settings, factors such as emotions, stress, or pain can affect a patient’s respiratory rate and CO2 production, adding additional layers of complexity not captured by the simulator (51). Furthermore, while we believe the waveform alterations and decreases in ETCO_2_ seen during CPAP in the HPS trials are consistent with what would be observed in human subjects, these results must be validated in clinical settings with actual patients to confirm their generalizability. The high flow rate of CPAP, which likely diluted exhaled CO_2_ and blunted the waveforms in the simulator, is inherent to CPAP therapy itself and would likely cause similar effects in human patients. However, human trials are necessary to verify this assumption and account for patient-specific factors.

Another limitation of this study is related to capnography sampling. This study primarily utilized nasal sampling with the CCA and oro-nasal sampling with the NC. In clinical practice, patients undergoing CPAP often breathe through their mouths, particularly in cases of respiratory distress, which could influence the capnography readings (32). The HPS does not differentiate between nasal and oral breathing, which limits the ability to analyze how different breathing patterns affect capnography measurements in humans. Future studies should investigate whether nasal, oral, or combined sampling methods provide the most accurate ETCO_2_ readings during CPAP in clinical settings.

While our study demonstrated significant changes in capnography waveforms and ETCO_2_ during CPAP administration, this does not imply that capnography is an ineffective tool for patients receiving CPAP. Capnography can still provide valuable insights into a patient’s ventilation status, but healthcare providers must be mindful of the limitations posed by high flow rates and CO_2_ dilution.

## Conclusions

Our study developed a novel CPAP-compatible capnography adapter (CCA) to address the issue of the nasal cannula (NC) disrupting the CPAP mask seal. Testing showed that both the CCA and NC provided accurate respiration rate measurements, but CPAP administration consistently reduced ETCO_2_, likely due to CO_2_ dilution from high flow rates. These findings suggest that current capnography protocols may not reliably reflect ventilation during CPAP therapy and that CPAP compromises capnogram morphology. Key next steps include conducting controlled clinical trials to evaluate these changes in human patients, particularly in prehospital and emergency care settings where capnography is widely used. These trials should assess the reproducibility of our findings across different patient populations and clinical conditions, such as COPD, asthma, and heart failure. Additionally, research should explore modifications to capnography sampling methods and equipment, such as alternative sampling locations or improved mask sealing, to mitigate CO_2_ dilution. Mainstream capnography may also warrant investigation as an alternative to sidestream methods commonly used in prehospital settings. Further, developing algorithms or software that adjust capnography readings for CPAP-related artifacts is another important avenue for future work. Machine learning models could potentially detect and compensate for waveform blunting and CO_2_ dilution, improving the diagnostic utility of capnography during CPAP. Studies combining capnography data with other physiological metrics, such as pulse oximetry, may provide more robust methods for assessing ventilation.

## Data Availability

All data produced in the present study are available upon reasonable request to the authors.

## Acknowledgments

The authors thank Steve Wilson for generously providing equipment that was vital for data collection. This work was funded by the Abrams Scholarship from the UNC/NCSU Joint Department of Biomedical Engineering, as well as significant in-kind support from the UNC School of Nursing Simulation Center, UNC/NCSU Joint Department of Biomedical Engineering, and Durham Technical Community College EMS Education program.

## Author Contributions

S.S. formed the idea, conducted the literature search, and designed the study; G.H., D.H, K.D., J.G., and I.R. provided technical feedback; S.S. and G.H. collected data; S.S. and I.R. prepared the manuscript; all authors reviewed, edited, and approved the manuscript.

## Declaration of Interests

The authors declare no competing interests.

## Abbreviations

EMS: Emergency Medical Services
CO_2_: Carbon Dioxide
CPAP: Continuous Positive Airway Pressure
ETCO_2_: End-Tidal Carbon Dioxide (mmHg)
PaCO_2_: Partial Pressure of Arterial Carbon Dioxide (mmHg)
SpO_2_: Blood Oxygen Saturation
RR: Respiration Rate (breaths/min)
NC: Nasal-Oral Cannula
CCA: CPAP-Capnography Adapter
MAP: Mean Arterial Pressure
HPS: Human Patient Simulator
PEEP: Positive End Expiratory Pressure

## Supporting Information

**Figure S1.**
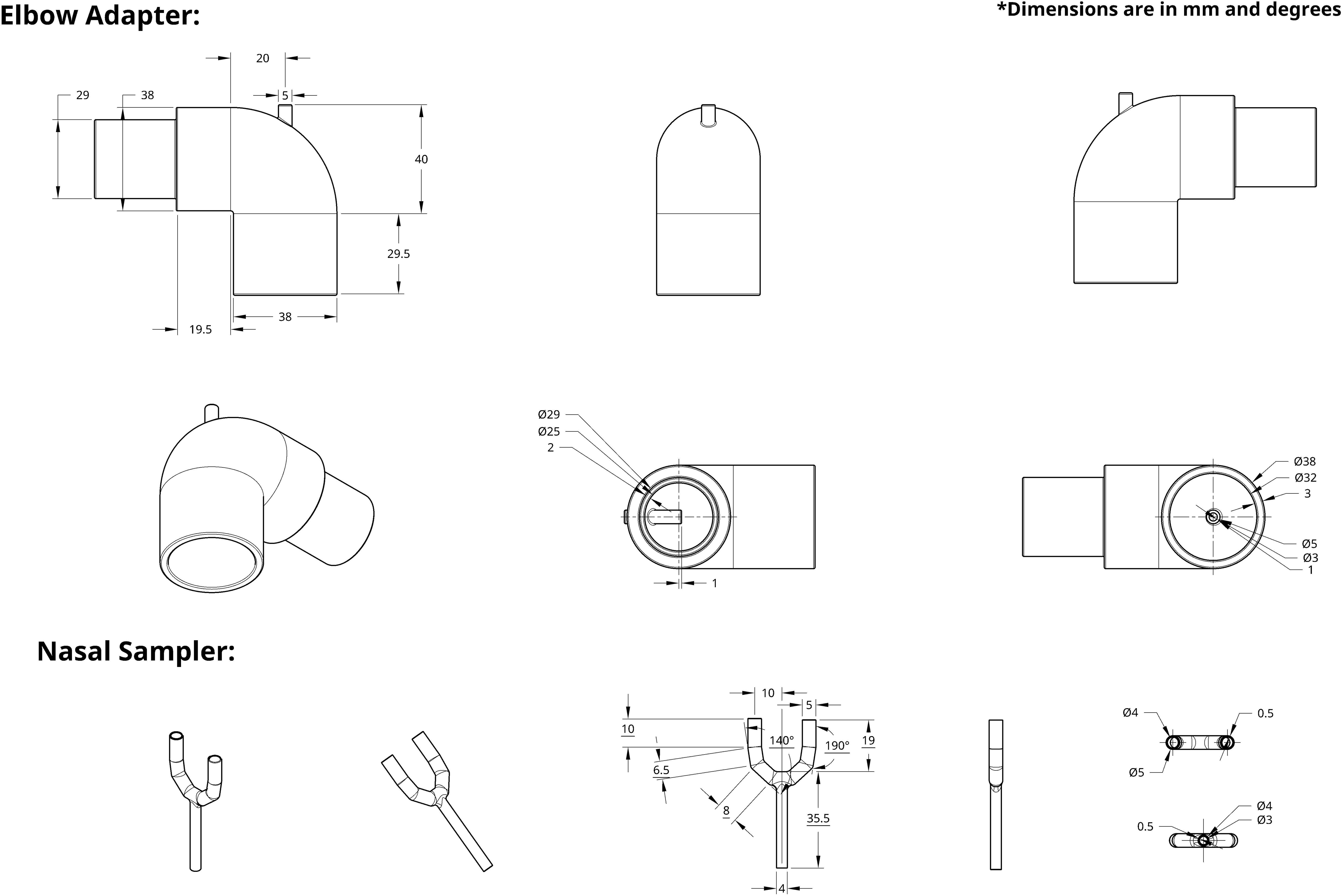
Design iterations of the CPAP-Capnography Adapter (CCA). (A) The first prototype of the CCA, showing an area with excess cured resin obstructing the internal diameter of the nasal prong. (B) The first prototype attached to CapnoLine® tubing and a CPAP mask; the exit point of the CapnoLine® tubing is labeled. (C) The second prototype of the CCA, showing the internal guide tube added to prevent kinking. (D) The nasal sampler of the CCA printed with Formlabs Clear V4 resin, with damage visible on the right nasal prong. CCA = CPAP-capnography adapter.

**Figure S2.**
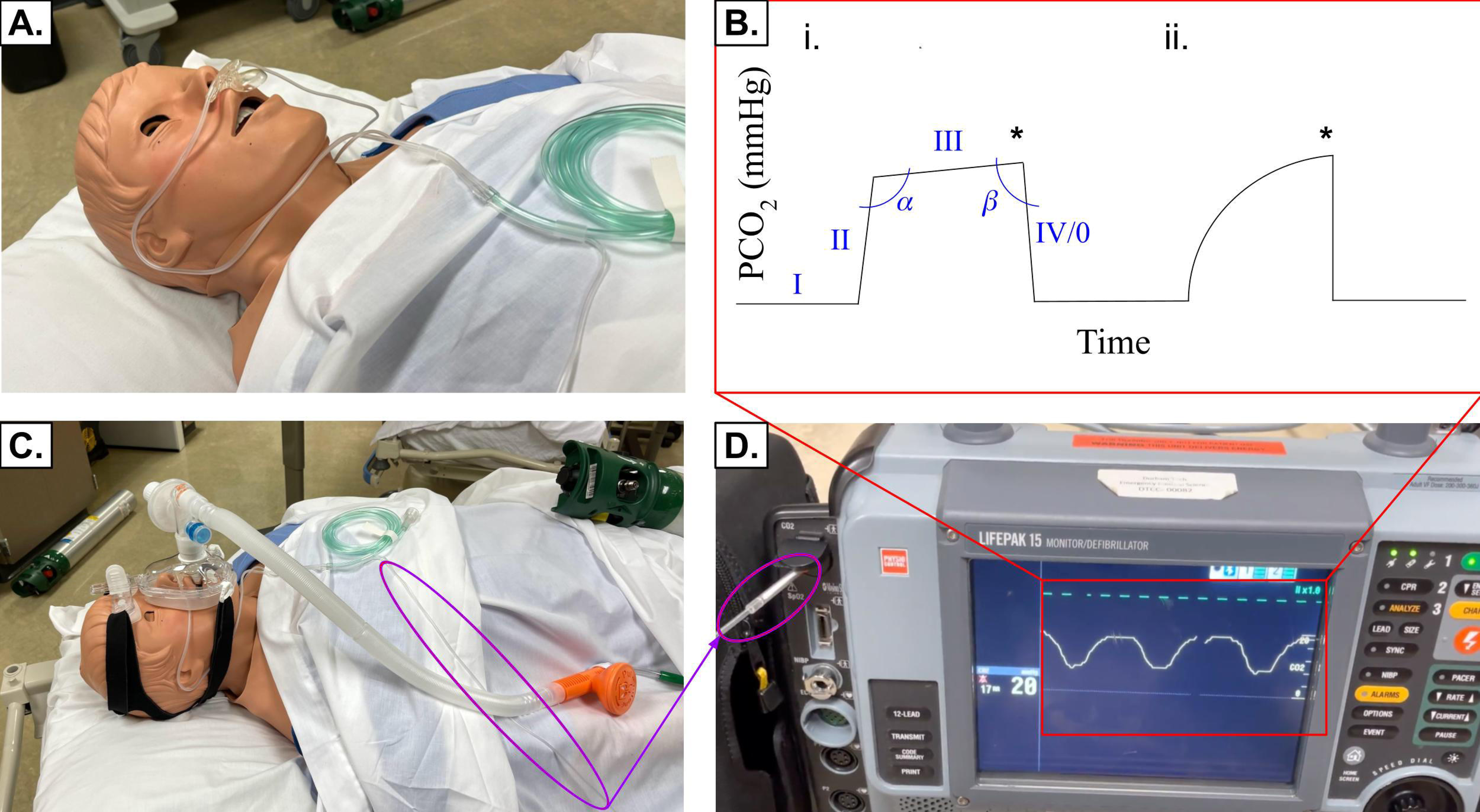
Representative capnography waveforms across all experimental groups for patient “Stan D. Ardman II” (Healthy Male). Representative waveforms from all experimental groups are shown. The consistency of the waveforms within each trial is highlighted. Capnography samplers and mask conditions are labeled. NC = naso-oral cannula, CCA = CPAP-capnography adapter, O_2_ = oxygen gas, CPAP = continuous positive airway pressure.

## Notes

### Competing Interest Statement

The authors have declared no competing interest.

### Funding Statement

This study did not receive any specific funding.

### Summary of Updates

To better reflect the scope and contribution of the study, we have revised the title, abstract, introduction, and discussion to focus on the device design and in vitro evaluation. We have also added a detailed description of the iterative development and testing of the CCA. The limitations section has been expanded to clearly delineate the constraints of mannequin-based research, and the implications for translation to human patients. Figures and captions have been revised for clarity and consistency.

